# Resting State fMRI Speech, Language, and Executive Function Network Connectivity in Children with and without Listening Difficulties

**DOI:** 10.1101/2021.10.21.21265296

**Authors:** Julia C. Hoyda, Hannah J. Stewart, Jennifer Vannest, David R. Moore

**Affiliations:** University of Cincinnati; University College London; Cincinnati Children’s Hospital and Medical Center; University of Manchester

## Abstract

Listening Difficulties (LiD) are characterized by a child having reported issues with listening despite exhibiting normal hearing thresholds. LiD can often overlap with other developmental disorders, including speech and language disorders, and involve similar higher-order auditory processing. This study used resting-state functional MRI to examine functional brain networks associated with receptive and expressive speech and language and executive function in children with LiD and typically developing (TD) peers (average age of 10 years).

We examined differences in region-of-interest (ROI)-to-ROI functional connectivity between: (1) the LiD group and the TD group and (2) within the LiD group, those participants who had seen a Speech-Language Pathologist and those who had not. The latter comparison was examined as a way of comparing children with and without speech and language disorders. Connections that differed between groups were analyzed for correlations with behavioral test data.

The results showed functional connectivity differences between the LiD group and TD group in the executive function network and trends in the speech perception network. Differences were also found in the executive network between those LiD participants who had seen an SLP and those who had not.

Several of these connectivity differences, particularly frontal-striatal connections, correlated with performance on behavioral tests: including tests that measure attention, executive function, and episodic memory, as well as speech, vocabulary, and sentence structure.

The results of this study suggest that differences in functional connectivity in brain networks associated with speech perception and executive function may underlie and contribute to listening difficulties.

## Introduction

Listening Difficulties (LiD) are often diagnosed when a child has reported issues with listening despite exhibiting normal hearing (decibel) thresholds (Moore et al. 2018). Listening, the active counterpart to hearing, encompasses multiple levels of processes, both neurological and physiological in nature from the inner ear to higher-order processing in the cortex.

Symptoms of LiD were initially observed as the consequences of lesions in the central auditory nervous system (CANS, Bocca et al. 1954), but more recently LiD has been characterized as any listening problem symptoms (not due to hearing loss) that may be caused by any one or combination of the following: attention deficits, auditory processing deficits, speech and language deficits, or other cognitive deficits (Moore 2018, Musiek et al. 2018). Because of the large overlap of these processes, one child with one set of symptoms may be given different diagnoses by different clinicians based on the vantage of their expertise, which may make treatment decisions challenging. In short, deficits in each of these areas may have similar behavioral manifestations.

Therefore, disentangling these different aspects of LiD proves very challenging. Although studies have attempted to use a variety of behavioral tasks to differentiate these processes (Gokula et al. 2019, Ferguson et al. 2011, Dawes & Bishop 2010, Sharma et al. 2009), any task that involves the perception of an auditory stimulus, including the most basic tone discrimination, is going to involve memory, attention, language (if only to understand the instructions), and executive function (Dillon & Cameron 2021). This becomes even more complex when speech and language are introduced into the task itself.

Therefore, these tasks may be able to inform us about how well a child with LiD performs compared to typically developing (TD) peers, but do not provide us with information about which processes are responsible for the lower performance. In other words, children who have LiD may perform uniformly on behavioral/psychometric measures, but have different neurological deficits underpinning those performances.

LiD has a high comorbidity with other developmental disorders, including speech and language disorders (de Wit et al. 2017). Sharma, et al. (2009) examined the overlap of diagnoses of Auditory Processing Disorder (APD), language impairment (LI), and reading difficulties (RD), finding that 47% of their participants had deficits in all three categories, whereas only 4% only had deficits unique to APD. In a testing battery by Dawes & Bishop (2010), APD and dyslexia were compared; they found no performance differences and that up to half of the children with APD could also fit the descriptions of dyslexia or specific language impairment (SLI) diagnoses. Ferguson et al. (2011) compared children with APD and SLI to TD children on various verbal and nonverbal tasks (such as digit span, sentence intelligibility, and nonsense word repetition) and found no differences in performance other than that the TD children performed better than both APD and SLI groups. They stated, “…[our finding] does not support the commonly held assumption that children with APD have specific difficulties listening in noise. It is suggested that poor attention may underlie these reported listening difficulties, although further evidence is required to support this” (p. 225). It is clear that difficulties with speech and language have a high degree of overlap with LiD but there may be differences in the interaction with attention and executive function.

Neuroimaging may allow us to better differentiate the neural mechanisms that contribute to LiD (Salimpoor et al. 2013, Schmithorst et al. 2011, Barton et al. 2012). Evidence for a high degree of interactivity between non-auditory, peripheral areas of the cortex and the auditory cortex (Hackett 2011, He & Yu 2010, Schofield 2010, Moore 2012) supports this idea. It can be said that higher-level cognitive processes regulate bottom-up primary auditory processes and the two meet in the middle in the auditory cortex (Moore & Hunter 2013). How these neurological mechanisms contribute to LiD is largely unknown; LiD may be underpinned by dysfunction in one or more contributing neural networks.

Prior analysis of resting state fMRI found significant connectivity differences between children with LiD and typically developing (TD) peers in regions supporting the processing of speech (Stewart et al., submitted). However, this analysis did not distinguish which aspects of speech or higher-order functioning contributed to the differences found. Therefore, our study dug deeper to understand more about these potential neural connectivity differences. Specifically, in this study, we used fMRI to examine resting state network connectivity in networks associated with expressive and receptive speech and language, as well as executive function in children with LiD compared to their TD peers.

We expect that those neurological networks supporting speech and language will show large connectivity differences between the LiD and TD groups. Specifically, those networks involved in speech and language *perception* would exhibit larger differences between the LiD and TD group than *production* networks, since LiD is manifested more clearly in perception tasks. We then also expected to find significant difference in executive function networks between the groups. To further examine the relationship between LiD and speech-language difficulties, we split the LiD group into those who had seen a speech-language pathologist (SLP), and those who had not, as a way of differentiating those with and without potential language disorders. Consequently, we expect to find connectivity differences between these two subgroups, specifically in the speech and language networks. Lastly, due to the roles of attention and cognitive function involved in listening, we expect to see connectivity differences between groups in the executive network.

## Methods

### Participants

This study included 81 children (52 male; mean age 10.01 years, SD 1.98) that were recruited as part of a larger study at CCHMC (Moore, et al. 2018) of over 1100 children for APD assessment. Children were recruited via questionnaires sent to their parents or caregivers, which included the Evaluation of Children’s Listening and Processing Skills (ECLiPS), and other relevant medical and demographic data. This recruitment grew over the course of the study to include social and digital media messages as well as print advertising at the hospital. Children with LiD versus typically developing children (TD) were categorized by the ECLiPS questionnaire, which is an inventory survey for caregivers to assess their child’s listening and hearing skills on a five-point Likert scale. A total standardized score of ≥ 5 was categorized as TD; and scores < 5, or a diagnosis of APD, were categorized as LiD. This identified 42 children as LiD and the remaining 39 as TD. All children participated in an audiology screening and exhibited normal thresholds of <25 dB HL at all octave-interval frequencies from 0.25 – 8 kHz in both ears. The children’s audiometric function and behavioral responses are reported elsewhere (Petley et al. in prep).

As a way of identifying those participants with LiD who may have a history of speech and language problems, we also divided the group based on parent-reported history of having seen a Speech-Language Pathologist (SLP). Of those participants in the LiD group, 26 were reported as having seen a Speech-Language Pathologist and 15 had not (1 exclusion due to questionnaire error).

### Parent report measures and behavioral tests

All participants completed the Children’s Communication Checklist-2 (CCC-2) is a parent or caregiver questionnaire that rates and scores the child’s skills in speech, semantics, syntax, language, social language, and more. It produces a norm-referenced score and an age-corrected general composite score (GCC). The NIH toolbox (Weintraub et al. 2013) is a test battery completed on a tablet covering several aspects of cognition including attention, memory, executive function, vocabulary, etc. Four subtests of the NIH toolbox were administered:

1. *Picture Vocabulary*: In the PV test, participants are shown four pictures and presented with an auditory stimulus. They must choose which picture best matches the meaning of the word.
2. *Flanker Inhibitory Control and Attention Test*: The FT tests inhibition/attention. Participants see a directional target with a stimulus on either side of the target. They must respond with the direction of the target (left or right) regardless of which side the side stimulus is on.
3. *Dimensional Change Card Sort test*: The DCCS measures attention and cognitive flexibility via rule switching. Participants must sort cards according to shape or color, which varies with each trial.
4. *Picture Sequence Memory test*: The PS measures episodic memory. Participants observe pairs of drawn objects/activities and an audio phrase. Sequences range from 6 to 18 pairs, and participants must remember pairs after two learning trials.

Participants were also administered the Listening in Spatialized Noise – Sentences (LISN-S, Cameron & Dillon 2007, 2008, 2009), which tests a participant’s ability to hear and recall target sentences despite distracting/competing sentences. These target and distraction sentences were presented at either the same or different locations around the participant (0 and/or 90 degrees). A minimum of 22 trials and a maximum of 30 trials were completed, stopping testing at SEM <1. Signal to noise ratio (SNR) increased or decreased with each trial depending on the participant’s correct or incorrect responses. 79 participants (40 LiD, all 39 TD) completed the LISN-S.

### Functional Magnetic Resonance Imaging

All children underwent a 5-minute resting state fMRI (TR/TE = 2000/30ms, voxel size = 2.5×2.5×3.5 mm, 39 axial slices, 150 volumes) in a 3T Philips Ingenia scanner with a 64-channel head coil. Children were told to hold still and focus on a fixation cross in the middle of a screen. All participants were awake and not sedated. The children also completed a T1-weighted anatomical scan (TR/TE = 8.1/3.7 ms, FOV = 25.6×25.6×16.0 cm, matrix = 256 × 256, and slice thickness = 1mm). Each child’s scans were performed in a single scanning session at Cincinnati Children’s Hospital.

## Data Analysis

### Regions of Interest (ROIs) maps

Regions of Interest (ROIs) were chosen from *Neurosynth*.*org* (an online meta-analysis of fMRI cortical activation studies, Yarkoni et al. 2011). In order to capture networks underlying multiple aspects of speech and language function, we chose four sets of ROIs using search terms ‘speech production’, ‘speech perception’, ‘naming’ (to capture expressive language), and ‘language comprehension’ (to capture receptive language). In addition, we examined an executive function network using the search term ‘executive’.

In order to construct networks of parcels of approximately equal anatomic size, maps from Neurosynth were parcellated using the ADHD200 Atlas (Bellec et al. 2017). Parcels 8 voxels or fewer in size were removed from analysis. The number of parcels in each network resulted as follows: ‘speech perception’ yielded 43 ROIs, ‘speech production’ 82, ‘language comprehension’ 54, ‘naming’ 56, and ‘executive’ 32. A representative slice view overlay of each ROI category can be seen in **Figure 10**.

### Resting state fMRI ROI-to-ROI analysis

rs-fMRI data was pre-processed using a standard pipeline (realignment, slice-timing correction, outlier identification, segmentation, normalization, and smoothing) in the Conn toolbox (Whitfield-Gabrieli and Nieto-Castanon 2012). Data is realigned by each scan being aligned to a referenced with the first image of the session. Slice-timing correction adjusts any temporal abnormalities or misalignments in the data by making sure the acquisition time matches throughout the scan. Outlier identification identifies data with potential as potential outliers due to either motion artifact or excessive BOLD signal. Segmentation and normalization occur when the data are aligned to the MNI brain space and segmented anatomically by matter type. Finally, functional smoothing is used to reduce variability and increase the BOLD signal-to-noise ratio (CONN toolbox 2021).

Pairwise functional temporal connectivity was examined within each set of ROIs by taking the average time course of all voxels in each ROI. Using Fisher’s *z*-transformation, *r* values were made into *z* values. These *z* values were used in group comparisons within each ROI network and in correlations with behavioral test scores.

### Group comparisons

Pairwise connectivity within each ROI network was examined for between-group comparisons. Two sets of group comparisons were made: (1) the LiD vs. TD groups and (2) within the LiD group, those participants who had seen a Speech-Language Pathologist (SLP) and those who had not (LiDSLP+ vs. LiDSLP-).

These connectivity group comparisons were made using Conn toolbox. Correction for multiple comparisons was applied using false discovery rate (Benjamin & Hochberg 1995). Group differences in connectivity results were examined at a threshold at p<0.05, corrected. To examine trends in the results, we also report results at a threshold of p<0.1 corrected.

### Relationships between connectivity and behavioral scores

Within those pairwise connections that differed between groups, we examined the relationship between connectivity and behavioral test scores using a Pearson correlation, to better understand how these connections related to children’s behavioral performance. The data were examined across all participants and thresholded at p<0.05.

## Results

### Behavioral Results

Children with LiD performed more poorly than their TD peers on all behavioral tests (see Table 1). However, among the children with LiD, there were no differences in performance between those children that had seen a Speech-Language Pathologist (SLP+) and those that had not (SLP-, all p>0.10). There was a trend toward children who had seen an SLP performing slightly lower on the LISN-S, though this was not significant p=0.085.

**Table 1.** Mean parent-report and behavioral scores in children with LiD and TD peers.

| Test | Group | n | Mean | SD | t | p |
| --- | --- | --- | --- | --- | --- | --- |
| CCC-2 | LiD | 42 | 82 | 14.98 | 10.701 | <0.001 |
|  | TD | 39 | 111.59 | 8.908 |  |  |
| NIH PV | LiD | 39 | 98.171 | 12.763 | 5.424 | <0.001 |
|  | TD | 38 | 114.864 | 14.224 |  |  |
| NIH FT | LiD | 39 | 89.287 | 9.9 | 3.952 | <0.001 |
|  | TD | 39 | 101.31 | 16.219 |  |  |
| NIH DCCS | LiD | 39 | 87.708 | 12.929 | 5.688 | <0.001 |
|  | TD | 39 | 105.292 | 14.34 |  |  |
| NIH PS | LiD | 36 | 94.05 | 19.842 | 4.646 | <0.001 |
|  | TD | 39 | 114.448 | 18.183 |  |  |
| LiSN-S | LiD | 40 | 0.198 | 1.2 | 0.243 | 0.809 |
|  | TD | 39 | 0.256 | 0.937 |  |  |

### Functional Connectivity - Group comparisons

#### TD vs. LiD

There were no differences in connectivity between the LiD and TD groups in the ‘speech production’, ‘naming’, or ‘language comprehension’ networks. However, four connections in the ‘speech perception’ network showed a trend of decreased connectivity in children with LiD compared to TD children (**Figure 2**, p<0.1 corrected). A trend toward reduced connectivity in the LiD group was observed between hemispheres and within the left hemisphere, including sections of the superior temporal gyrus (STG), Heschl’s gyrus (HG), Rolandic operculum (RO) and inferior postcentral gyrus. Five connections in the same network showed a trend toward increased connectivity in children with LiD compared to the TD group (**Figure 2**, p<0.1).

**Figure 1:**
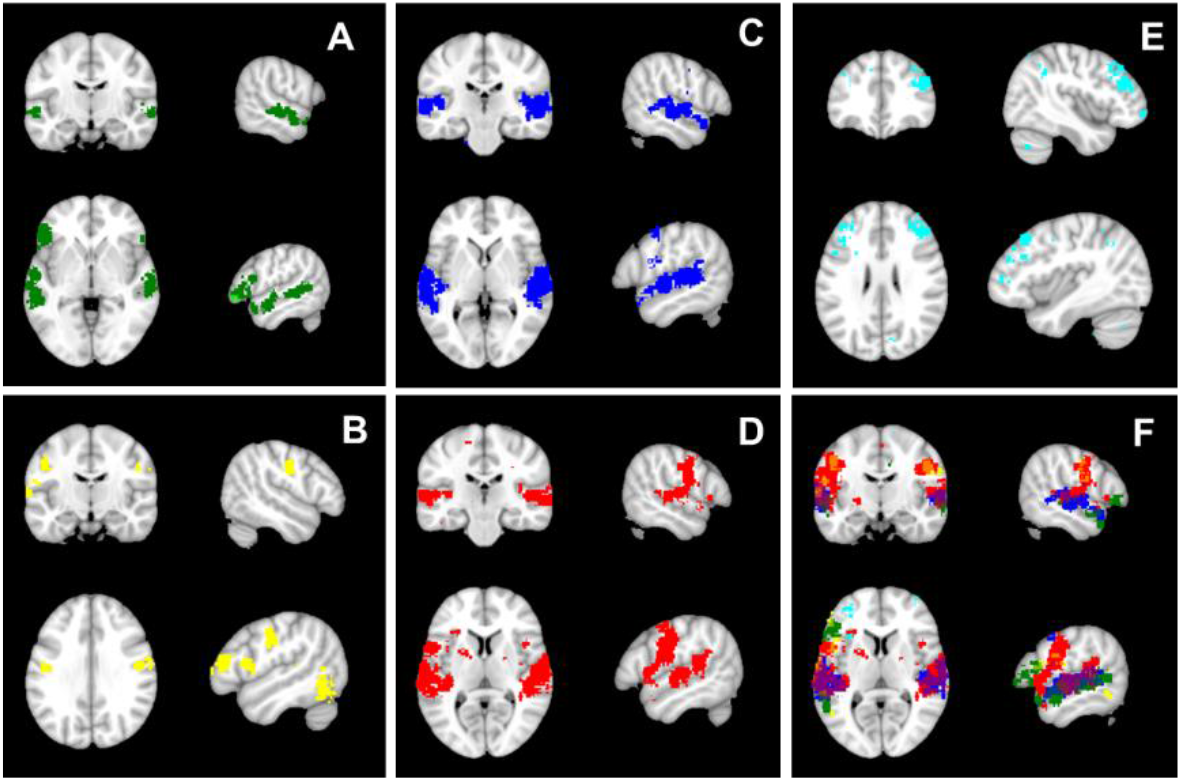
Sample slices of each set of ROIs. (A) Language Comprehension, (B) Naming, (C) Speech Production, (D) Speech Perception, (E) Executive, and (F) overlay of all ROIs.

**Figure 2:**
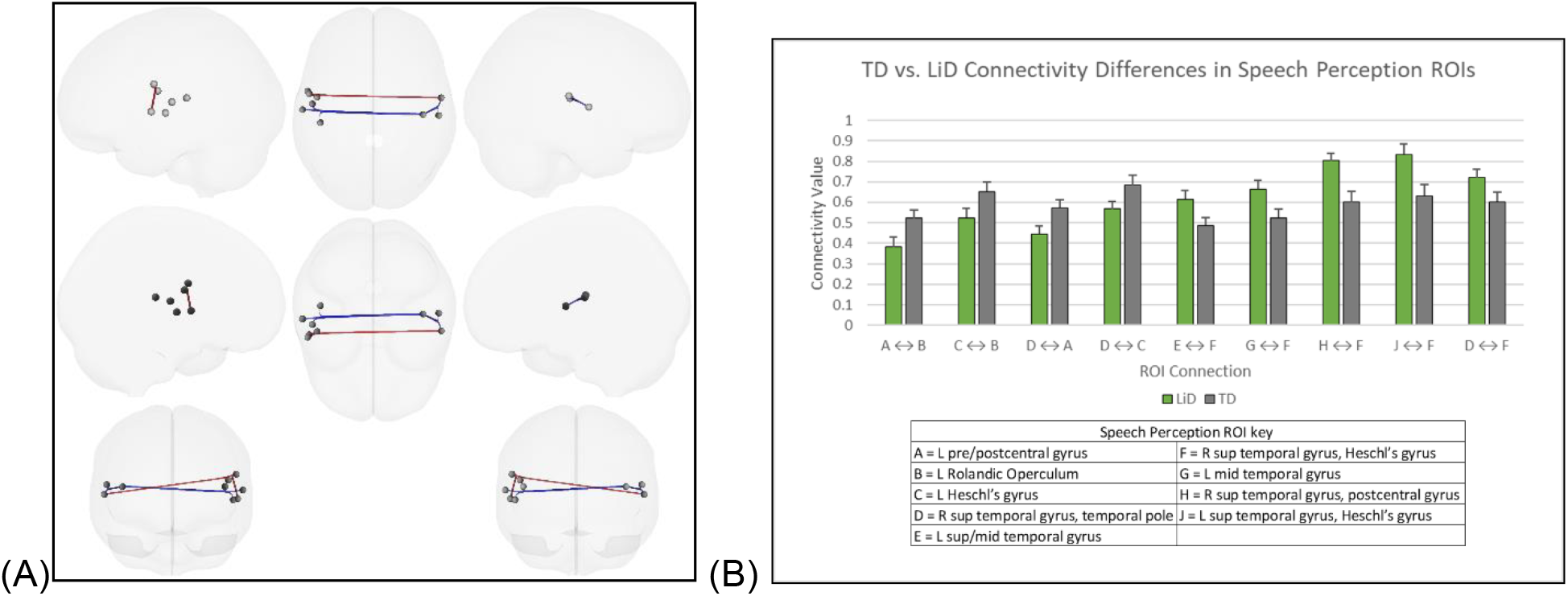
(*A*) 3D brain rendering of TD vs. LiD contrast in Speech Perception ROIs. Red is where TD connectivity>LiD connectivity; Blue is where LiD>TD. (*B*) The connectivity values for each significant connection from (A).

Increased connectivity in the LiD group was observed between hemispheres and within the right hemisphere, all of which were connections involving the right STG/HG. Connectivity with an ROI at the junction of the right STG, RO and HG was both reduced and increased in children with LiD depending on the connection (see **Figure 2**).

Within the executive function network, there was significantly decreased connectivity within the left hemisphere in children with LiD compared to TD children. Three of the five connectivity differences where the LiD group had lower connectivity than the TD group, occurred between the left caudate and left midfrontal gyrus. Children with LiD had significantly increased connectivity compared to TD children in relation to one region in the left inferior parietal/angular gyrus (in connections with the left superior midfrontal gyrus and with the left inferior parietal gyrus). (**Figure 3**, p<0.05, corrected)

**Figure 3:**
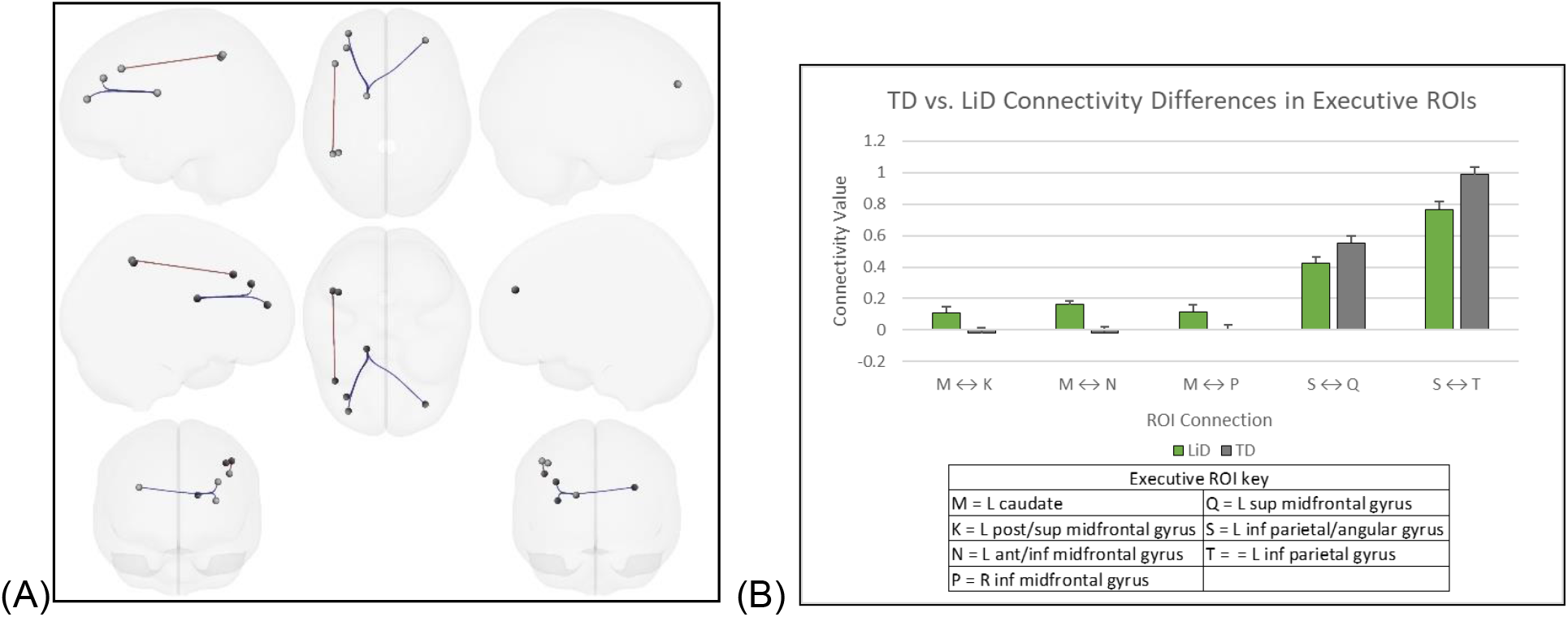
(*A*) 3D brain rendering of TD vs. LiD contrast in Executive ROIs. Red is where TD connectivity>LiD connectivity; Blue is where LiD>TD. (*B*) The connectivity values for each significant connection from (A), ROI key can be found in **Table 2**. Negative values indicate anti-correlation or negative functional connectivity (NFC).

#### LiD(SLP+) vs. LiD(SLP-)

There were no significant differences in connectivity between SLP+ and SLP-in the ‘speech production’, ‘naming’, ‘speech perception’, or ‘language comprehension’ networks. Within the executive network, children with LiD who had seen an SLP (SLP+) had significantly decreased connectivity, five total connections (both across hemispheres and within the left hemisphere) compared to their peers who had not seen an SLP (SLP+ > SLP-). These differences appeared in connections between ROIs including the left midfrontal gyrus, left insula and inferior frontal gyrus pars triangularis, and the right midfrontal gyrus (**Figure 4**, p<0.05, corrected). There were no significant connectivity differences in the reverse direction (SLP+ > SLP-).

**Figure 4:**
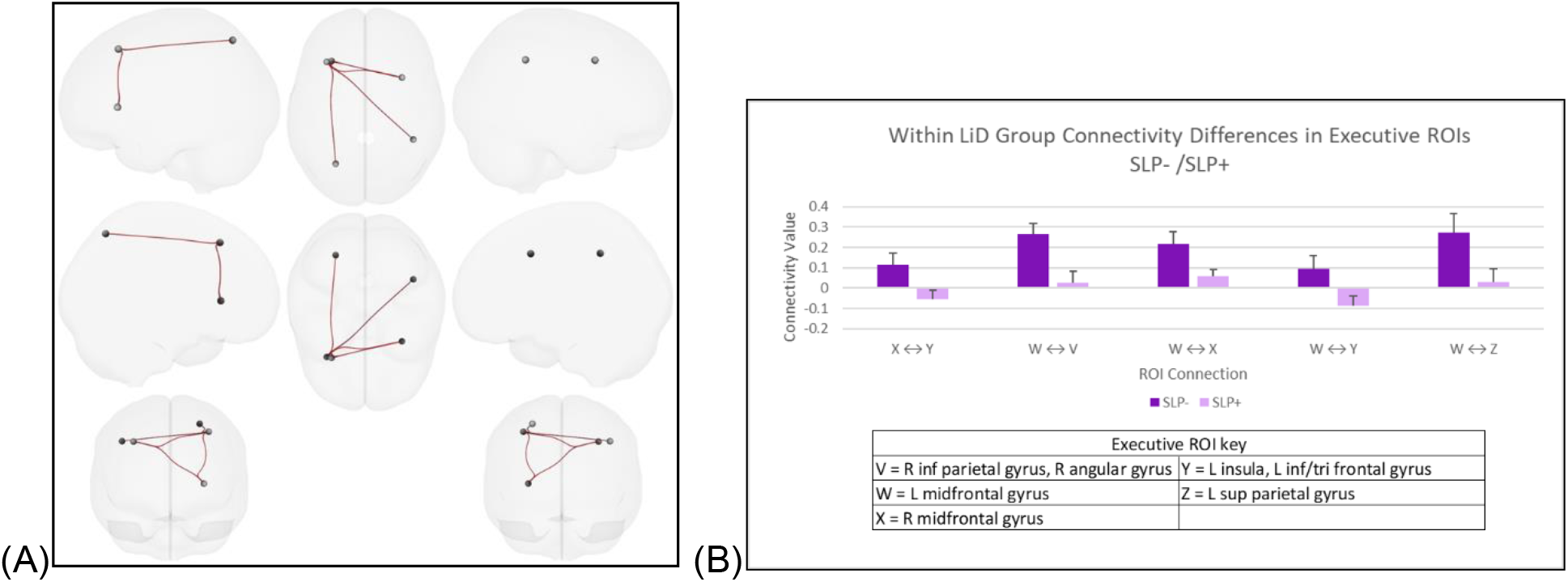
(*A*) 3D brain rendering of LiDs with and without report of seeing a Speech Language Pathologist (SLP+/-) in Executive ROIs. Red is where SLP-connectivity>SLP+ connectivity. (*B*) The connectivity values for each significant connection from (A). Like in Figure 3, NFC occurred, this time in the LiD(SLP+) group.

### Relationships between connectivity and behavioral scores

As noted above, five pairwise connections within the executive network differed significantly (p<0.05) between groups. These five connections were as follows: (1) left caudate to left posterior/superior midfrontal gyrus, (2) left caudate to left anterior/inferior midfrontal gyrus, (3) left caudate to right inferior midfrontal gyrus, (4) left superior midfrontal gyrus to left inferior parietal/angular gyrus, and (5) left inferior parietal/angular gyrus to left inferior parietal gyrus. In these five connections, we examined the relationship between connectivity and behavioral test scores using Pearson correlation.

#### CCC-2

Two connections had significant correlations with CCC-2 GCC scores (**Figure 5**). Connectivity between the left caudate and left anterior-inferior midfrontal gyrus correlated negatively with CCC-2 scores (p = 0.001, r = -0.358). Connectivity between the left angular gyrus and left inferior parietal gyrus correlated positively with CCC-2 scores (p < 0.001, r = 0.387).

**Figure 5:**
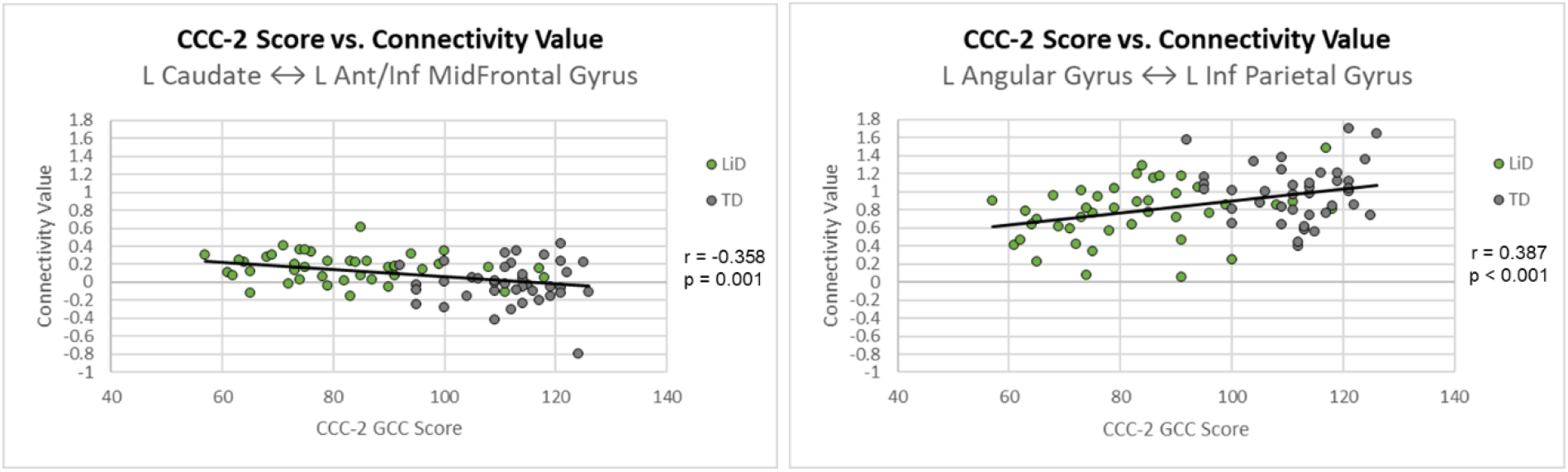
Correlations of Children’s Communication Checklist General Composite scores and executive function network connectivity values.

#### NIH toolbox, PV

Connectivity between the left caudate to left anterior-inferior midfrontal gyrus correlated negatively with the Picture Vocabulary test scores of the NIH toolbox (**Figure 6**, p = 0.01, r = -0.292).

**Figure 6:**
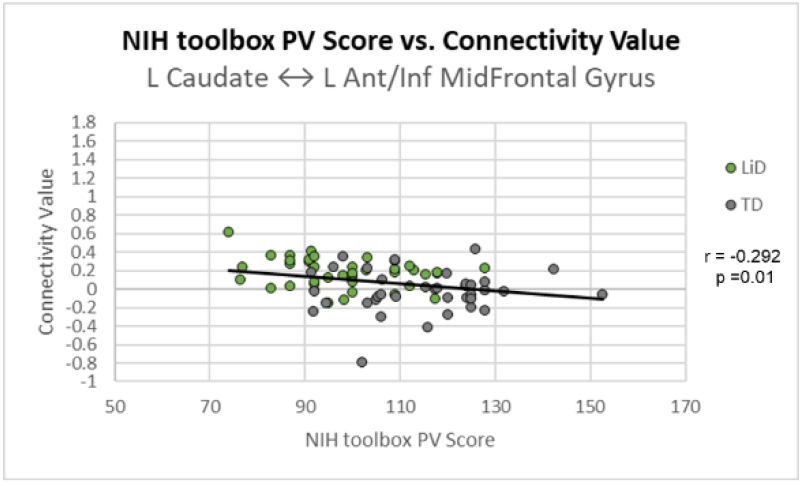
Correlation of NIH toolbox Picture Vocabulary scores and executive function network connectivity values.

#### NIH toolbox, FT

Connectivity between the left caudate to left anterior-inferior midfrontal gyrus correlated negatively with Flanker Test scores of the NIH toolbox (**Figure 7**, p = 0.026, r = - 0.252).

**Figure 7:**
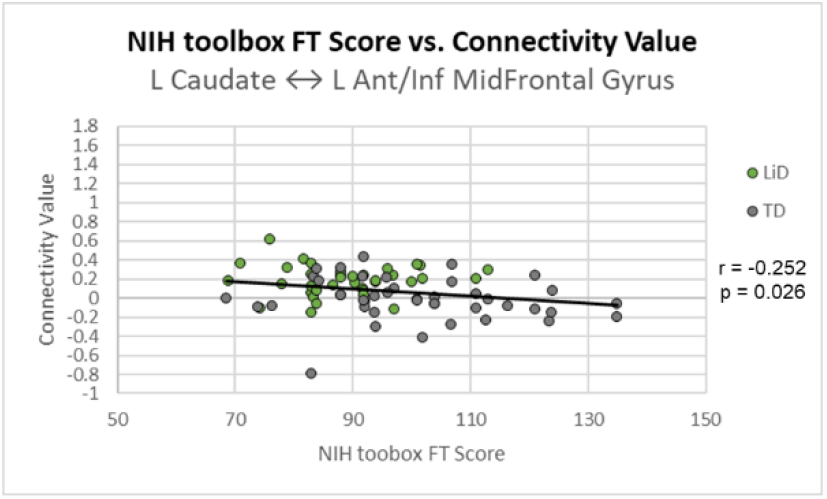
Correlation of NIH toolbox Flanker Test scores and executive function network connectivity values.

#### NIH toolbox, DCCS

Three connections correlated with the Dimensional Change Card Sorting task (**Figure 8**). Connectivity between the left caudate to left anterior-inferior midfrontal gyrus correlated negatively with DCCS scores (p < 0.001, r = -0.439). Connectivity between the left inferior parietal/angular gyrus and left superior midfrontal gyrus, and left inferior parietal gyrus correlated positively with DCCS scores (p = 0.026, r = 0.251; p = 0.041, r = 0.232, respectively)

**Figure 8:**
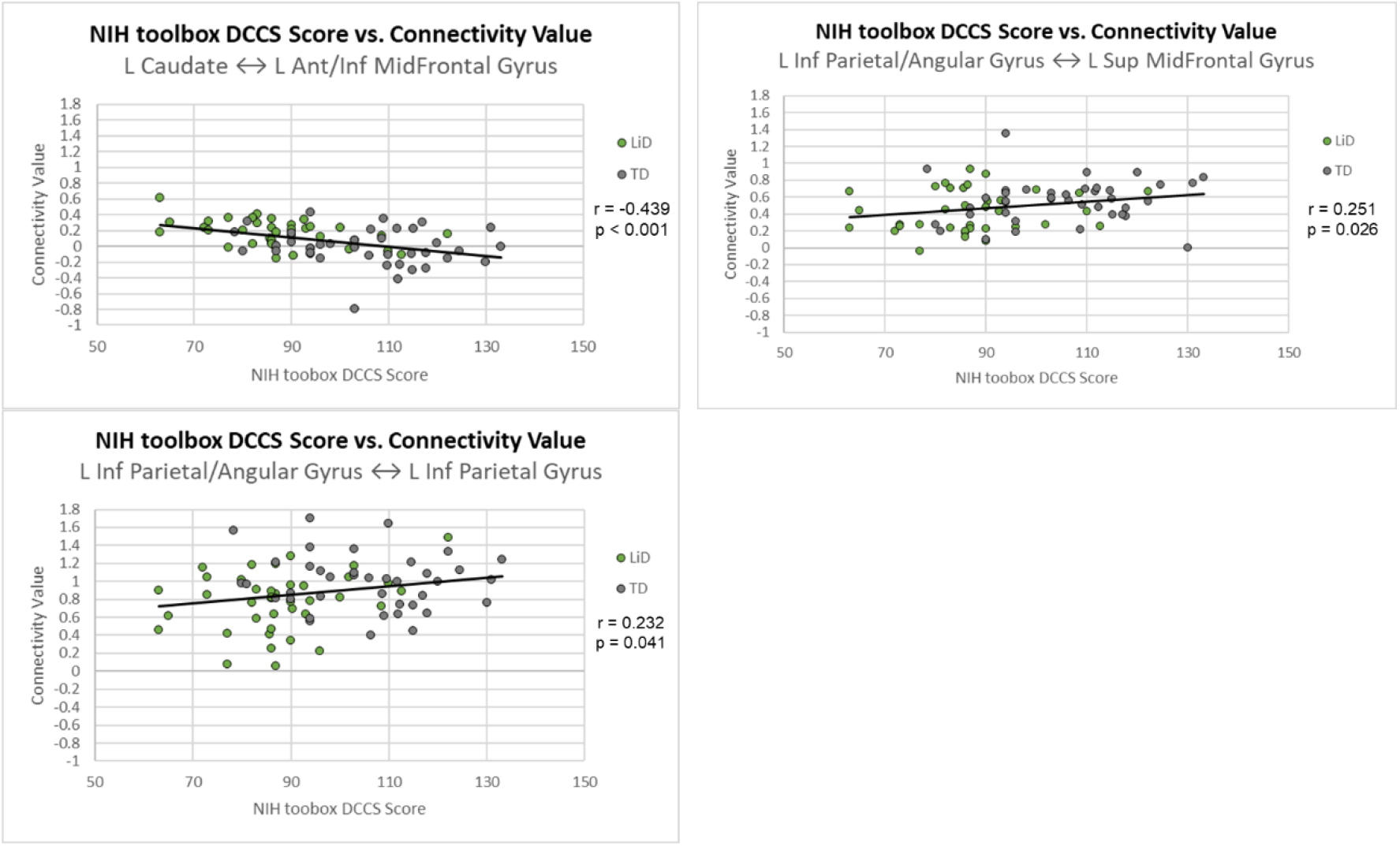
Correlations of NIH toolbox Dimensional Change Card Sorting Task scores and executive function network connectivity values.

#### NIH toolbox, PS

Two connections correlated with Picture Sequence Memory scores of the NIH toolbox (**Figure 9**). Connectivity between the left caudate to left anterior-inferior midfrontal gyrus correlated negatively with PS scores (p = 0.032 r = -0.247). Connectivity between the left parietal/angular gyrus and left inferior parietal gyrus (p = 0.006, r = 0.316) correlated positively with PS scores.

**Figure 9:**
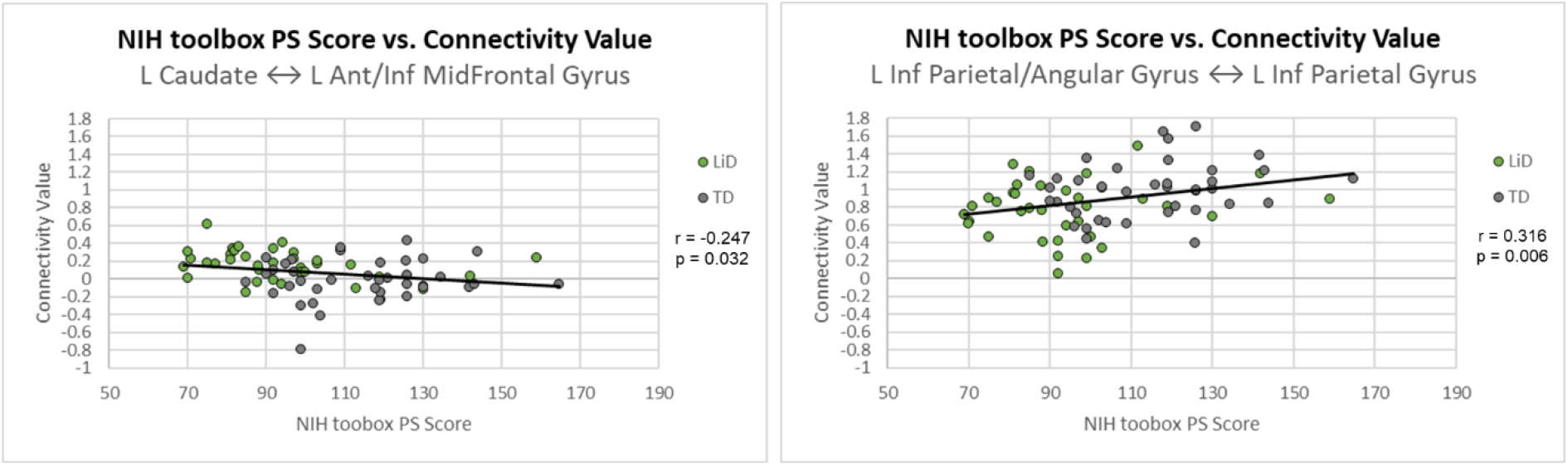
Correlations of NIH toolbox Picture Sequence Memory task scores and executive function network connectivity values.

#### LISN-S

No significant correlations were found with executive network connections.

## Discussion and Conclusions

We used resting-state functional MRI connectivity to examine potential differences in *a-priori* speech, language and executive function networks between children with Listening Difficulties (LiD) and their typically-developing peers. Behaviorally, we found that children with LiD performed more poorly than their peers based on parent report of language skills, as well as tests of vocabulary and executive function. Because deficits in the processing of auditory information is a hallmark of LiD, we hypothesized that the speech perception network would show the strongest group differences among the four speech/language networks we examined. However, the expressive and receptive speech and language networks did not show differences in connectivity between the LiD and TD groups. There was, however, a trend within the speech perception network that indicated hyperconnectivity within right hemisphere regions and cross-hemispheric regions in the children with LiD, suggesting that their speech perception networks may be atypical.

Importantly, the greatest differences in functional connectivity between children with LiD and TD peers were within the executive network, where there were five significant connectivity differences; children with LiD showed hypoconnectivity in compared to TD children in three connections each involving an ROI in the left caudate. In contrast, they exhibited hyperconnectivity in two connections involving an ROI in the left inferior parietal/angular gyrus.

These differences in functional connectivity also corresponded directly to poorer behavioral performance. In particular, scores on the CCC-2 and behavioral tests were significantly negatively correlated with connectivity between the left caudate and left anterior/inferior midfrontal gyrus (which was overall shown to be hyperconnected in children with LiD). The more poorly an individual child performed in terms of overall language skills, vocabulary and executive function, the **greater** the connectivity in this connection.

Conversely, there were other connections where we found an overall pattern of hypoconnectivity in children with LiD, and poorer performance was associated with reduced connectivity. This was observed in connections within the left parietal lobe. Specifically, connectivity between the left angular and left inferior parietal gyrus was reduced in children with lower CCC-2 scores. Reduced connectivity between the left inferior parietal/angular gyrus and the left inferior parietal gyrus was associated with poorer with Picture Sequencing and Dimensional Change Card Sort scores on the NIH Toolbox.

All of these findings suggest that changes in functional connectivity in the executive network may contribute to the range of behavioral deficits found in children with listening difficulties. These findings are consistent with current literature indicating that disruptions in the executive network are indicators for executive dysfunction and attention problems including ADHD. A study by De Simoni et al. (2018), found altered caudate connectivity to be correlated with executive dysfunction in adults with TBI. Similar findings are present in preschool children (Hawkey et al. 2018) where both EEG and fMRI showed disrupted patterns of connectivity in executive networks predicted ADHD. Another EEG study (Han & Chan 2017) showed that children with ASD who had executive dysfunction exhibited disordered connectivity in the fronto-parietal network. Other studies, which define the caudate as being part of the “cool” network in ADHD (networks responsible for cognitive flexibility, sustained attention, etc.) (Cubillo et al. 2012), show that hyperactivity and restlessness in adults correlate with hyperconnectivity in the caudate and putamen (Sörös et al. 2019) and that reduced connectivity in fronto-striatal regions may contribute to reduced cognitive flexibility (Vaghi et al. 2017) but higher connectivity in those regions correlates with severity of executive dysfunction in ADHD (Li et al. 2014). Because we observed group connectivity differences in many of the same regions in the executive network, it is likely that attention and executive dysfunction play larger roles in LiD than speech and language networks than we anticipated.

Among children with LiD, we used the child’s history of having seen a Speech-Language Pathologist as a way of identifying those participants with LiD who may have more significant speech and language problems. Interestingly, at time of testing for this study, the SLP+ and SLP-groups did not differ in their CCC-2 scores or performance on any other testing. One explanation for this may be that the speech and language issues of the SLP+ group may have either been previously resolved through speech/language therapy or not severe enough when compared to their SLP-peers to see a difference in performance. There were also no differences in speech/language networks. However, hypoconnectivity was found among children with LiD who had seen an SLP versus those who had not<u>, particularly in connections among the</u> left midfrontal gyrus, left insula and inferior frontal gyrus (pars triangularis), and right midfrontal gyrus: this was a different set of regions and connections than those where differences were observed in the LiD group as a whole.

Overall, our results have shown that resting state fMRI is a sensitive method for examine the neural mechanisms underlying LiD. This avoids the need to create a task that successfully isolates just one of the aspects or skills needed for auditory processing. With both bottom-up and top-down processes at play, the neural mechanisms of LiD can stem from anywhere within the processing pathway, so examination of broad brain networks is key in this approach. In addition, due to the high comorbidity of LiD with speech and language disorders such as SLI and dyslexia, it was crucial to examine connectivity that speech and language neural networks exhibit in LiD. Likewise, other higher-order functions like attention and memory serve important roles in listening; therefore, we also examined neural network connectivity in the executive network. Our findings of group differences in connectivity in the executive network, and their relationship to behavioral performance, suggest that these mechanisms play a greater role in LiD and associated language and executive deficits more than previously thought. Specifically, the neural underpinnings of LiD symptoms may lie more in disrupted higher-order processes like attention and learning and may be less tied to disruptions in language processing or decoding information about the auditory stimuli. Further work is needed to better characterize the interaction of listening difficulties with speech and language disorders across development

## Data Availability

All data produced in the present study are available upon reasonable request to the authors

## Bibliography

Moore, D. R. (2018). Guest Editorial: Auditory Processing Disorder. Ear & Hearing, 39(4), 617–620. Doi: 10.1097/AUD.0000000000000582

E. Bocca, C. Calearo & V. Cassinari (1954) A New Method for Testing Hearing in Temporal Lobe Tumours: Preliminary report, Acta Oto-Laryngologica, 44:3, 219–221, DOI: 10.3109/00016485409128700

Musiek, Frank E; Iliadou, Vasiliki (Vivian); Chermak, Gail D.; Bamiou, Doris-Eva; Rawool, Vishakha Waman; Ptok, Martin; Purdy, Suzanne; Jutras, Benoît; Moncrieff, Deborah; Stokkereit Mattsson, Tone; Ferre, Jeanane M.; Fox, Cydney; Grech, Helen; Geffner, Donna; Hedjever, Mladen; Bellis, Teri James; Nimatoudis, Ioannis; Eleftheriadis, Nikos; Pedersen, Ellen Raben; Weihing, Jeffrey; Guillory, Lisa; Madell, Jane R.; Hurley, Annette; Whitelaw, Gail M.; Schochat, Eliane; Spyridakou, Chrysa; Sidiras, Christos; Thai-Van, Hung; Kostopoulou, Anastasia; Veuillet, Evelyne; Keith, Bill; Mountjoy, Alyson; Koohi, Nehzat; Sirimanna, Tony; Lau, Carol; Cone, Barbara; Kiese-Himmel, Christiane; Abramson, Maria; & Raghunathrao, Rangasayee. (2018) Letter to the Editor: An Affront to Scientific Inquiry Re: Moore, D. R. (2018) Editorial: Auditory Processing Disorder, Ear Hear, 39, 617–620, Ear and Hearing: November/December 2018 - Volume 39 - Issue 6 - p 1236-1242, DOI: 10.1097/AUD.0000000000000644

Gokula, R., Sharma, M., Cupples, L., & Valderrama, J. T. (2019). Comorbidity of auditory processing, attention, and memory in children with word reading difficulties. Frontiers in Psychology, 10(OCT), 1–15. https://doi.org/10.3389/fpsyg.2019.02383

Ferguson, M. A., Hall, R. L., Riley, A., & Moore, D. R. (2011). Communication, listening, cognitive and speech perception skills in children with auditory processing disorder (APD) or specific language impairment (SLI). Journal of Speech, Language, and Hearing Research, 54(1), 211+. http://dx.doi.org.proxy.libraries.uc.edu.uc.idm.oclc.org/10.1044/1092-4388(2010/09-0167)

Dawes, P., & Bishop, D. V. M. (2010). Psychometric profile of children with auditory processing disorder and children with dyslexia. Archives of Disease in Childhood, 95(6), 432–436. https://doi.org/10.1136/adc.2009.170118

Sharma, Mridula; Purdy, Suzanne C; & Kelly, Andrea S. (2009) Comorbidity of Auditory Processing, Language, and Reading Disorders. Journal of Speech, Language, and Hearing Research; Jun 2009; 52, 706–722

Dillon, H., & Cameron, S. (2021). Separating the Causes of Listening Difficulties in Children. Ear & Hearing, Publish Ah, 1–12. https://doi.org/10.1097/aud.0000000000001069

de Wit, E., van Dijk, P., Hanekamp, S., Visser-Bochane, M., Steenbergen, B., van der Schans, C. P., & Luinge, M. R. (2017). Same or Different: The Overlap Between Children With Auditory Processing Disorders and Children With Other Developmental Disorders: A Systematic Review. Ear & Hearing, 39(1), 1–19. Doi: 10.1097/AUD.0000000000000479

Barton B, Venezia JH, Saberi K, Hickok G, Brewer AA. (2012) Orthogonal acoustic dimensions define auditory field maps in human cortex. Proc Nat Acad Sci USA. 109: 20738–43.

Salimpoor VN, van den Bosch I, Kovacevic N, McIntosh AR, Dagher A, Zatorre RJ. (2013) Interactions between the nucleus accumbens and auditory cortices predict music reward value. Science (New York, NY). 340: 216–9.

Schmithorst VJ, Holland SK, Plante E. (2011) Diffusion tensor imaging reveals white matter microstructure correlations with auditory processing ability. Ear Hear. 32: 156–67.

Hackett, T. A. (2011). Information flow in the auditory cortical network. Hearing Research, 271(1–2), 133–146.

He, J., & Yu, Y. (2010). Role of descending control in the auditory pathway. In A. Rees & A. R. Palmer (Eds.), The oxford handbook of auditory science: The auditory brain (pp. 247–268). New York: Oxford University Press.

Schofield, B. R. (2010). Structural organization of the descending auditory pathway. In A. Rees & A. R. Palmer (Eds.), The oxford handbook of auditory science: The auditory brain (pp. 43–64). New York: Oxford UP.

Moore, DR. (2012). Listening difficulties in children: Bottom-up and top-down contributions. Journal of Communication Disorders, 45, 411–418.

Moore, D. R., & Hunter, L. L. (2013). Auditory processing disorder (APD) in children: A marker of neurodevelopmental syndrome. Hearing, Balance & Communication, 11(3), 160–167. https://doi-org.uc.idm.oclc.org/10.3109/21695717.2013.821756

Stewart, H. J.; Cash, E. K.; Hunter, L. L.; Maloney, T.; Vannest, J.; & Moore, D. R. (submitted) Speech Cortical Activation and Connectivity in Typically Developing Children and Those with Listening Difficulties. Health & Amp; Medicine Week, NewsRX LLC, p. 6863. https://go.exlibris.link/PDKTLgyy

Petley, L., Hunter, L., Motlagh Zadeh, L., Stewart, H., Sloat, N., Perdew, A., Lin, L. & Moore, D. (in prep). Listening Difficulties in Children With Normal Audiograms. Ear and Hearing, Publish Ahead of Print, doi: 10.1097/AUD.0000000000001076.

Weintraub S, Dikmen SS, Heaton RK, Tulsky DS, Zelazo PD, Bauer PJ, Gershon RC. Cognition assessment using the NIH Toolbox. (2013) Neurology. 80(11 Suppl. 3) S54–S64.

Cameron, S. & Dillon, H. (2007). Development of the Listening in Spatialized Noise-Sentences Test (LISN-S). Ear and Hearing, 28 (2), 196–211. doi: 10.1097/AUD.0b013e318031267f.

Cameron, S., & Dillon, H. (2008). The listening in spatialized noise-sentences test (LISN-S): comparison to the prototype LISN and results from children with either a suspected (central) auditory processing disorder or a confirmed language disorder. Journal of the American Academy of Audiology, 19(5), 377–391. https://doi.org/10.3766/jaaa.19.5.2

Cameron, S. (Designer), & Dillon, H. (Designer). (2009). Listening in Spatialized Noise – Sentences test (LISN-S). Software, Phonak Communications AG. https://www.phonakpro.com/au/en/resources/fitting-and-tests/lisn-s-test/overview-lisn-s.html

Yarkoni, T., Poldrack, R. A., Nichols, T. E., Van Essen, D. C., & Wager, T. D. (2011). Large-scale automated synthesis of human functional neuroimaging data. Nature Methods, 8(8), 665–670. Doi: 10.138/NMETH.1635

Bellec, P., Chu, C., Chouinard-Decorte, F., Benhajali, Y., Margulies, D. S., & Craddock, R. C. (2017). The neuro bureau ADHD-200 preprocessed repository. Neuroimage, 144, 275–286. doi: 10.1016/j.neuroimage.2016.06.034

Whitfield-Gabrieli S, Nieto-Castanon A. (2012). Conn: a functional connectivity toolbox for correlated and anticorrelated brain networks. Brain Connect, 2(3), 125–41. Doi: 10.1089/brain.2012.0073

CONN toolbox - Preprocessing pipeline. (n.d.). http://Web.conn-Toolbox.org. Retrieved October 1, 2021, from https://web.conn-toolbox.org/fmri-methods/preprocessing-pipeline#h.p_jISegaTEhy9a

Benjamin, Y. & Hochberg, Y. (1995) Controlling the False Discovery Rate: a Practical and Powerful Approach to Multiple Testing. J.R. Statist. Soc. B. 57(1), 289–300.

De Simoni, S., Jenkins, P. O., Bourke, N. J., Fleminger, J. J., Hellyer, P. J., Jolly, A. E., Patel, M. C., Cole, J. H., Leech, R., & Sharp, D. J. (2018). Altered caudate connectivity is associated with executive dysfunction after traumatic brain injury. Brain, 141(1), 148–164. https://doi.org/10.1093/brain/awx309

Hawkey, E. J., Tillman, R., Luby, J. L., & Barch, D. M. (2018). Preschool Executive Function Predicts Childhood Resting-State Functional Connectivity and Attention-Deficit/Hyperactivity Disorder and Depression. Biological Psychiatry: Cognitive Neuroscience and Neuroimaging, 3(11), 927–936. https://doi.org/10.1016/j.bpsc.2018.06.011

Han, Y. M. Y., & Chan, A. S. (2017). Disordered cortical connectivity underlies the executive function deficits in children with autism spectrum disorders. Research in Developmental Disabilities, 61, 19–31. https://doi.org/10.1016/j.ridd.2016.12.010

Cubillo, A., Halari, R., Smith, A., Taylor, E., & Rubia, K. (2012). A review of fronto-striatal and fronto-cortical brain abnormalities in children and adults with Attention Deficit Hyperactivity Disorder (ADHD) and new evidence for dysfunction in adults with ADHD during motivation and attention. Cortex, 48(2), 194–215. https://doi.org/10.1016/j.cortex.2011.04.007

Sörös, P., Hoxhaj, E., Borel, P., Sadohara, C., Feige, B., Matthies, S., Müller, H. H. O., Bachmann, K., Schulze, M., & Philipsen, A. (2019). Hyperactivity/restlessness is associated with increased functional connectivity in adults with ADHD: A dimensional analysis of resting state fMRI. BMC Psychiatry, 19(1), 1–11. https://doi.org/10.1186/s12888-019-2031-9

Vaghi, M. M., Vértes, P. E., Kitzbichler, M. G., Apergis-Schoute, A. M., van der Flier, F. E., Fineberg, N. A., Sule, A., Zaman, R., Voon, V., Kundu, P., Bullmore, E. T., & Robbins, T. W. (2017). Specific Frontostriatal Circuits for Impaired Cognitive Flexibility and Goal-Directed Planning in Obsessive-Compulsive Disorder: Evidence From Resting-State Functional Connectivity. Biological Psychiatry, 81(8), 708–717. https://doi.org/10.1016/j.biopsych.2016.08.009

Li, F., He, N., Li, Y., Chen, L., Huang, X., Lui, S., Guo, L., Kemp, G. J., & Gong, Q. (2014). Intrinsic Brain Abnormalities in Attention Deficit Hyperactivity. Radiology, 272(2), 514–523.

